# A SARS-CoV-2 Nucleocapsid Variant that Affects Antigen Test Performance

**DOI:** 10.1101/2021.05.05.21256527

**Authors:** Lori Bourassa, Garrett A. Perchetti, Quynh Phung, Michelle J. Lin, Margaret G. Mills, Pavitra Roychoudhury, Kimberly G. Harmon, Jonathan C. Reed, Alexander L. Greninger

## Abstract

More than one year into a global pandemic, SARS-CoV-2 is now defined by a variety of rapidly evolving variant lineages. Several FDA authorized molecular diagnostic tests have been impacted by viral variation, while no reports of viral variation affecting antigen test performance have occurred to date. While determining the analytical sensitivity of the Quidel Sofia SARS Antigen FIA test (Sofia 2), we uncovered a high viral load specimen that repeatedly tested negative by this antigen test. Whole genome sequencing of the specimen uncovered two mutations, T205I and D399N, present in the nucleocapsid protein of the isolate. All six SARS-CoV-2 positive clinical specimens available in our laboratory with a D399N nucleocapsid mutation and C_T_ < 31 were not detected by the Sofia 2 but detected by the Abbott BinaxNOW COVID-19 Ag Card, while clinical specimens with the T205I mutation were detected by both assays. Testing of recombinant SARS-CoV-2 nucleocapsid with these variants demonstrated an approximate 1000-fold loss in sensitivity for the Quidel Sofia SARS Antigen FIA test associated with the D399N mutation, while the BinaxNOW and Quidel Quickvue SARS Antigen tests were unaffected by the mutation. The D399N nucleocapsid mutation has been relatively uncommon to date, appearing in only 0.02% of genomes worldwide at time of writing. Our results demonstrate how routine pathogen genomics can be integrated into the clinical microbiology laboratory to investigate diagnostic edge cases, as well as the importance of profiling antigenic diversity outside of the spike protein for SARS-CoV-2 diagnostics.

## Introduction

The second year of the SARS-CoV-2 pandemic has been marked by the emergence of a variety of lineages of the virus, several of which are associated with increased transmissibility and immune escape. These variants are generally marked by mutations in the spike protein, which undergoes the most rapid evolution in the coronaviruses due the continual need to escape humoral immunity (1–4). The effect of viral evolution has affected every aspect of the pandemic, from concerns over increased mortality to changes in therapeutic and vaccine efficacy and diagnostic assay performance (5, 6).

To date, the FDA has noted four molecular tests whose performance could be altered by SARS-CoV-2 evolution (7). Most famously, S gene target failures in the Thermo TaqPath assay mark an N-terminal domain deletion in the spike protein that helps define the B.1.1.7 lineage currently increasing in prevalence across the United States (8). Unlike molecular testing, the effect of SARS-CoV-2 variants on antigen testing is not well understood. No variant affecting antigen test performance has been described to date. Most SARS-CoV-2 antigen tests target the nucleocapsid protein since it is stably associated with the RNA in the virion and present at higher copy number than other viral structural proteins such as spike (9). Most evolutionary analyses in coronavirus have focused on spike protein, since it is responsible for receptor binding and entry and thus a constant target of neutralizing antibodies (4, 10). However, with more than 18 months of spread in humans, a number of coding changes have been found in the nucleocapsid protein (11). Here, we describe the characterization of an uncommon nucleocapsid variant that affects the performance of the Quidel Sofia SARS Antigen FIA antigen test run on the Sofia 2 platform.

## Materials and Methods

### Specimens and study design

Deidentified, residual patient nasopharyngeal (NP) samples submitted to the University of Washington Virology Laboratory, Seattle, WA for clinical diagnostic testing were used in this study. To minimize potential impact of different viral transport media on assay performance, only clinical samples collected in phosphate-buffered saline (PBS) were used for analytical sensitivity determination. This study was approved by the University of Washington Institutional Review Board.

### Quidel Sofia 2 analytical sensitivity determination and confirmation

To determine the limit of detection, we created a pooled positive specimen by equally mixing three residual nasopharyngeal (NP) samples collected in PBS with C_T_s of 23.7, 16.8 and 17.6 by a CDC-based laboratory developed test (12, 13). The pooled specimen was quantified using two methodologies: reverse transcription-droplet digital PCR (RT-ddPCR, Bio-Rad, Hercules, CA, USA) and qRT-PCR with the SARS-CoV-2 assay on the cobas 6800 (Roche, Basel, Switzerland) as previously described (14).

Serial 10-fold dilutions of the neat pooled positive sample were run in triplicate on the Sofia 2 antigen test. 50µL of each dilution was applied to sterile swabs provided with the Sofia 2 kit to simulate sample collection from the nares (15). The Quidel Reagent Solution was added to a reagent tube and the nasal swab was mixed with the kit reagents by rolling the swab several times and pressing the swab against the bottom of the tube as recommended by the manufacturer. After incubating for 1 minute at room temperature, approximately 120µL of the sample was pipetted into the test cassette. Each cassette was allowed to develop for 15 minutes and was read using the Sofia 2 instrument.

In addition to the pooled specimen, 58 deidentified residual NP samples collected in PBS that previously tested positive for SARS-CoV-2 on the Roche cobas 6800 with a E-gene C_T_ range 14.27-37.97 were tested on the Sofia 2 SARS Antigen FIA following the methods described above.

### Clinical specimen testing on Abbott BinaxNOW

Testing of clinical specimens with the Abbott BinaxNOW COVID-19 Ag Card was performed as described previously (15). Briefly, swabs provided with the kit were spiked with 50µL of clinical nasal specimens previously collected into 3mL of PBS (16). The spiked swab specimen was tested following the manufacturer’s instructions.

### Quantitative RT-PCR and viral load determination of clinical specimens

Viral load was determined by testing on the Roche cobas 6800 using the Seracare AccuPlex SARS-CoV-2 Verification Panel standards. SARS-CoV-2 E-gene calibrators of 1e3 (C_T_ 33.41), 1e4 (C_T_ 30.71), and 1e5 copies (C_T_ 27.43) were used to calculate -0.33 slope and 14.17 intercept from the Roche cobas 6800 platform. Copies per swab were calculated by dividing the copies/mL by 20, based on the 50µL of sample liquid used for rapid test.

### Whole genome sequencing and genome mining

SARS-CoV-2 whole genome sequencing was performed using the Swift Biosciences v2 or Illumina COVID-Seq amplicon tiling platforms as described previously (17). To determine descriptive epidemiology of the nucleocapsid D399N mutation, SARS-CoV-2 whole genomes were downloaded from GISAID on April 18, 2021 (18). Figures were generated with custom R code (github.com/greninger-lab/sars_cov2_antigen_test_n_gene), using ggplot (18), UpSetR (19), and PANGO lineages (20).

### Cloning of SARS-CoV-2 N gene wild-type and mutants

The ORF encoding N wild-type was amplified from a plasmid encoding N sequence based on NC_045512.2 (a gift from Michael Gale Jr lab) using the COVID-N-C-Strep-F and COVID-N-C-Strep-R primers. To obtain the ORF encoding N T205I/D399N, RNA was extracted from a previously sequenced clinical SARS-CoV-2 isolate that encodes the T205I/D399N mutations in N (WA-UW-62718; EPI_ISL_1366195) and converted to cDNA using Superscript IV (ThermoFisher Scientific, Waltham, MA, USA)) with COVID-N-C-Strep-R primer followed by PCR utilizing the same primer pair that amplified N wild-type gene. The N wild-type or N T205I/D399N amplicons were then cloned into a modified pcDNA4/TO vector with a C-terminal 2x Strep-Tag II using the In-Fusion HD cloning kit (Takara Bio, Kusatsu, Shiga, Japan). To obtain the construct encoding the N T205I amino acid change an amplicon encoding the first half of the N gene encoding T205I (made by PCR using primers COVID-N-C-Strep-F and COVID N nt864R with the N T205I/D399N cDNA template) and an amplicon encoding the second half of the N wild-type gene (made by PCR using primers COVID N nt864R and COVID-N-C-Strep-R with the N wild-type plasmid template), were cloned into the modified pcDNA4/TO vector with a C-terminal 2x Strep-Tag II using the In-Fusion HD cloning kit (Takara). The N D399N construct was made following the same strategy except by combining an amplicon encoding the first half of the N wild-type gene (made by PCR using primers COVID-N-C-Strep-F and COVID N nt864R with the N wild-type plasmid) and an amplicon encoding the second half of the N gene encoding D399N (made by PCR using primers COVID N nt864R and COVID-N-C-Strep-R with the N T205I/D399N cDNA template). The sequence of all plasmids was confirmed by Sanger sequencing (Genewiz, Inc.) and all primers are listed in Table S1. All PCR reactions were performed with CloneAmp Hi-Fi PCR premix (Takara).

### Cell culture, transfection, and cell lysis

293T cells were maintained in Dulbecco’s modified Eagle medium (DMEM) high glucose with 10% fetal bovine serum (FBS), 10 mM HEPES, penicillin and streptomycin. Cells were transfected with 1 µg of plasmid DNA using 3:1 ratio of PEI MAX (Polysciences, Inc. Warringston, PA, USA) in Opti-MEM. Cells were harvested 48 h after transfection by washing cells with PBS (with Ca^2+^ and Mg^2+^) followed by lysis in 1xLSM lysis buffer (20 mM HEPES pH 7.9, 14 mM KAc, 1 mM NaCl, 1 mM MgAcetate_2_, 0.3% NP40) with 1% protease inhibitor cocktail (Sigma-Aldrich, St. Louis, MO, USA). Cell lysates were clarified by centrifugation at 200xg for 10 min at 4°C, followed by a second clarification by centrifugation at 21,100g for 1 min at 4°C. Lysates were then stored at -80°C prior to analysis.

### Confirming SARS-CoV-2 N expression and estimating concentration of SARS-CoV-2 N in lysates

Total protein in 293T cell lysates was measured using the Pierce BCA protein assay kit (ThermoFisher) and the samples were normalized to 125 µg/mL in sample buffer. To estimate the concentration of N in cell lysates a titration of recombinant his-tagged N protein (ThermoFisher) was prepared ranging from 62.5 ng to 250 ng in sample buffer. Lysates (1.25✉μg per lane) and the titration of recombinant his-tagged N (ThermoFisher) were run on a 4-12% Bis-Tris sodium dodecyl sulfate (SDS)-polyacrylamide gel with morpholinepropanesulfonic acid (MOPS) running buffer (Invitrogen, Carlsbad, CA, USA) under reducing conditions. The samples were then transferred to a 0.45-μm nitrocellulose membrane using the XCell Blot II module (Invitrogen). Blotting was performed with 1:500 anti-SARS-CoV-2 Nucleocapsid Protein (E8R1L) mouse IgG_2a_ monoclonal antibody (Cell Signaling Technology, Danvers, MA, USA) followed by staining with 1:2,000 IRDye 800CW anti-mouse IgG secondary antibody (Li-Cor Biosciences, Lincoln, NE, USA). Blots were scanned on Sapphire Biomolecular Imager (Azure Biosystems, Dublin, CA, USA) and quantified using Image Studio Lite version 5.2.5 (Li-Cor).

To estimate the concentration of N in the lysate, the estimated nanograms of N quantified by western blot was divided by 10 to account for loading volume and then multiplied by dilution factor required to bring the lysates to 0.125 mg/mL (lysates were diluted 22.4-fold, 25.6-fold, 21.6-fold, and 28-fold for N WT, N T205I, N D399N, and N T205I/D399N, respectively). This resulted in a concentration of 336.2 ng/µL, 189.7 ng/µL, 403.9 ng/µL, 244.2 ng/µL for N WT, N T205I, N D399N, and N T205I/D399N, respectively. These values were used to calculate pg quantity of N going into the 1:10 dilution for N variant based on absorbing 50 µL diluted lysate for each antigen test.

### Abbott BinaxNOW, Quidel Sofia 2, and Quidel QuickVue testing of cell lysates

Lysates from 293T cells transfected with N wildtype, N T205I, N D399N, N T205I/D399N, or pcDNA4/TO vector were kept frozen at -80°C prior to use in antigen testing. Lysates were thawed and diluted in 1xLSM lysis buffer at the indicated ten-fold dilutions (from neat to 1:10^6^). Subsequent dilutions tested between 10-fold dilutions were either serially diluted from existing 10-fold dilutions or a new 10-fold dilution was made and retested. For each test, 50 µL of sample was transferred to 1.5 mL microcentrifuge tube and the material was absorbed to a kit specific swab. The swab was tested following the manufacturer’s instructions. BinaxNOW and Quidel QuickVue SARS Antigen Test results were independently evaluated by two individuals, and specimens were called negative if disagreement, inclusive of the 1:100,000 T205I BinaxNow, 1:100,000 WT QuickVue, 1:100,000 D399N QuickVue specimens.

## Results

### Quidel Sofia SARS Antigen FIA analytical sensitivity is approximately 54,900 copies/mL

To determine the analytical sensitivity of the Sofia SARS Antigen FIA on the Sofia 2 analyzer (referred to here as the Sofia 2 test), a stock pooled positive sample was determined to have a viral load of 8.66×10^6^ copies/mL by the Roche cobas SARS-CoV-2 assay and 1.10×10^7^ copies/mL by RT-ddPCR. Using ten-fold serial dilutions of the pooled sample tested in triplicate, the initial limit of detection (LoD) was determined to be between 1:10 and 1:100 (Table 1). Twenty additional 1:10 dilutions were tested to confirm the LoD. However, only 12/20 (60%) specimens of the 1:10 dilution were detected by the assay. Therefore, 1:9 dilutions were tested, with 20/20 (100%) samples detected. Quantification of the 1:9 dilution by the Roche cobas 6800 and RT-ddPCR estimated the Sofia 2 SARS Antigen FIA LoD between 9.62×10^5^ and 1.22×10^6^ virus copies/mL, respectively. To account for the lower sample volume used by the rapid antigen test compared to RT-PCR platforms (50 µL compared to 200-500 µL used by commercial RT-PCR platforms) a copies/swab and theoretical C_T_ calculation for the 50µL volume absorbed on the swab was included to better represent the LoD of the antigen test. The LoD of the Sofia 2 SARS antigen FIA was determined at 4.81 x10^4^ and 6.11×10^4^ virus copies/swab by cobas 6800 and RT-ddPCR, respectively, or an approximate theoretical qRT-PCR C_T_ of 29.

**Table 1.** Analytical sensitivity of Quidel SARS Antigen FIA test using dilution of a pooled positive

| **Sample Dilution** | **Initial Replicates Detected** | **Initial % Positive** | **Confirmatory Replicates Detected** | **Copies/mL (ddPCR)** | **Copies/swab (ddPCR)** | **Copies/mL (Cobas 6800)** | **Copies/swab (Cobas 6800)** |
| --- | --- | --- | --- | --- | --- | --- | --- |
| Neat | 3/3 | 100% |  | 1.10E+07 | 5.50E+05 | 8.66E+06 | 1.73E+05 |
| **1:9** | **3/3** | **100%** | **20/20** | **1.22E+06** | **6.11E+04** | **9.62E+05** | **4.81E+04** |
| **1:10** | **3/3** | **100%** | **12/20** | **1.10E+06** | **5.50E+04** | **8.66E+05** | **4.33E+04** |
| 1:20 | 0/3 | 0% |  | 5.50E+05 | 2.75E+04 | 4.33E+05 | 8.66E+03 |
| 1:30 | 0/3 | 0% |  | 3.67E+05 | 1.83E+04 | 2.89E+05 | 5.77E+03 |
| 1:40 | 0/3 | 0% |  | 2.75E+05 | 1.38E+04 | 2.16E+05 | 4.33E+03 |
| 1:50 | 0/3 | 0% |  | 2.20E+05 | 1.10E+04 | 1.73E+05 | 3.46E+03 |
| 1:100 | 0/3 | 0% |  | 1.10E+05 | 5.50E+03 | 8.66E+04 | 1.73E+03 |
| 1:1,000 | 0/3 | 0% |  | 1.10E+04 | 5.50E+02 | 8.66E+03 | 4.81E+01 |
| 1:10,000 | 0/3 | 0% |  | 1.10E+03 | 5.50E+01 | 8.66E+02 | 4.33E+01 |
| 1:100,000 | 0/3 | 0% |  | 1.10E+02 | 5.50E+00 | 8.66E+01 | 8.66E+00 |
| Pos Control | 6/6 | 100% |  |  |  |  |  |
| Neg Control | 0/6 | 0% |  |  |  |  |  |

To determine whether this analytical sensitivity was reflected in a broader set of specimens, we performed antigen testing on 58 deidentified residual NP swabs collected for clinical diagnostic testing that comprised a wide range of viral loads (C_T_ 14.27-37.97). Of the 23 samples included within the LoD established using the pooled specimen, 22/23 (95.7%) were detected by rapid testing. Three samples below the LoD (2.25×10^4^, 1.24×10^4^, 4.66×10^3^ copies/swab) were detected but none beyond 4,660 virus copies/swab (0/30), corresponding to a theoretical cobas 6800 C_T_ of 32.9, were detected (Figure 1, Table 2).

**Table 2.** Confirmation of Analytical Sensitivity of Quidel SARS Antigen FIA test on Sofia 2 analyzer using clinical specimens. Clinical specimens in PBS transport media were spiked onto swabs for antigen test, resulting in a calculated theoretical C_T_ value of how much virus was used as input for the antigen test.

| UW<br>Sample ID | Original<br>Roche<br>cobas C<sub>T</sub> | Original<br>Copies /<br>mL | Copies /<br>swab | Theoretical<br>Roche<br>cobas C<sub>T</sub> | Quidel<br>SOFIA 2<br>Antigen<br>Detection |
| --- | --- | --- | --- | --- | --- |
| PBS 21 | 14.24 | 2.67E+09 | 1.34E+08 | 19.6 | Positive |
| PBS 10 | 14.49 | 2.20E+09 | 1.10E+08 | 19.9 | Positive |
| PBS 2 | 15.12 | 1.36E+09 | 6.80E+07 | 20.5 | Positive |
| PBS 7 | 17.23 | 2.69E+08 | 1.35E+07 | 22.6 | Positive |
| PBS 33 | 17.41 | 2.34E+08 | 1.17E+07 | 22.8 | Positive |
| PBS 35 | 19.18 | 6.02E+07 | 3.01E+06 | 24.5 | Positive |
| PBS 32 | 19.19 | 5.98E+07 | 2.99E+06 | 24.5 | Positive |
| PBS 29 | 19.35 | 5.28E+07 | 2.64E+06 | 24.7 | Positive |
| PBS 38 | 19.8 | 3.74E+07 | 1.87E+06 | 25.2 | Positive |
| PBS 55 | 20.38 | 2.40E+07 | 1.20E+06 | 25.7 | Positive |
| PBS 15 | 20.5 | 2.19E+07 | 1.09E+06 | 25.8 | Positive |
| PBS 37 | 20.52 | 2.15E+07 | 1.08E+06 | 25.9 | Positive |
| PBS 53 | 21.01 | 1.48E+07 | 7.39E+05 | 26.4 | Positive |
| PBS 14 | 21.29 | 1.19E+07 | 5.96E+05 | 26.6 | Positive |
| PBS 45 | 21.3 | 1.18E+07 | 5.91E+05 | 26.6 | Positive |
| PBS 19 | 21.56 | 9.69E+06 | 4.84E+05 | 26.9 | Positive |
| PBS 52 | 21.58 | 9.54E+06 | 4.77E+05 | 26.9 | Positive |
| PBS 22 | 21.63 | 9.18E+06 | 4.59E+05 | 27.0 | Positive |
| PBS 26 | 22.52 | 4.64E+06 | 2.32E+05 | 27.9 | Positive |
| PBS 36 | 22.63 | 4.26E+06 | 2.13E+05 | 28.0 | Negative |
| PBS 34 | 22.66 | 4.16E+06 | 2.08E+05 | 28.0 | Positive |
| PBS 51 | 23.23 | 2.69E+06 | 1.34E+05 | 28.6 | Positive |
| PBS 25 | 24.22 | 1.26E+06 | 6.28E+04 | 29.6 | Positive |
| PBS 47 | 24.48 | 1.03E+06 | 5.15E+04 | 29.8 | Negative |
| PBS 27 | 25.56 | 4.49E+05 | 2.25E+04 | 30.9 | Positive |
| PBS 57 | 26.33 | 2.49E+05 | 1.24E+04 | 31.7 | Negative |
| PBS 28 | 26.35 | 2.45E+05 | 1.23E+04 | 31.7 | Positive |
| PBS 9 | 27.61 | 9.31E+04 | 4.66E+03 | 32.9 | Positive |
| PBS 12 | 28.13 | 6.25E+04 | 3.12E+03 | 33.5 | Negative |
| PBS 43 | 28.56 | 4.49E+04 | 2.25E+03 | 33.9 | Negative |
| PBS 23 | 29.14 | 2.88E+04 | 1.44E+03 | 34.5 | Negative |
| PBS 58 | 29.15 | 2.86E+04 | 1.43E+03 | 34.5 | Negative |
| PBS 30 | 29.3 | 2.54E+04 | 1.27E+03 | 34.6 | Negative |
| PBS 50 | 29.36 | 2.43E+04 | 1.22E+03 | 34.7 | Negative |
| PBS 8 | 29.61 | 2.01E+04 | 1.00E+03 | 34.9 | Negative |
| PBS 1 | 30.21 | 1.27E+04 | 6.33E+02 | 35.5 | Negative |
| PBS 11 | 30.26 | 1.22E+04 | 6.09E+02 | 35.6 | Negative |
| PBS 41 | 30.53 | 9.90E+03 | 4.95E+02 | 35.8 | Negative |
| PBS 31 | 30.79 | 8.11E+03 | 4.05E+02 | 36.1 | Negative |
| PBS 3 | 30.83 | 7.86E+03 | 3.93E+02 | 36.1 | Negative |
| PBS 16 | 31.43 | 4.96E+03 | 2.48E+02 | 36.7 | Negative |
| PBS 48 | 31.74 | 3.91E+03 | 1.95E+02 | 37.1 | Negative |
| PBS 18 | 31.82 | 3.68E+03 | 1.84E+02 | 37.1 | Negative |
| PBS 44 | 31.87 | 3.54E+03 | 1.77E+02 | 37.2 | Negative |
| PBS 5 | 32.55 | 2.10E+03 | 1.05E+02 | 37.9 | Negative |
| PBS 46 | 32.67 | 1.91E+03 | 9.57E+01 | 38.0 | Negative |
| PBS 17 | 32.79 | 1.75E+03 | 8.73E+01 | 38.1 | Negative |
| PBS 42 | 33.2 | 1.27E+03 | 6.37E+01 | 38.5 | Negative |
| PBS 54 | 33.2 | 1.27E+03 | 6.37E+01 | 38.5 | Negative |
| PBS 49 | 33.58 | 9.52E+02 | 4.76E+01 | 38.9 | Negative |
| PBS 39 | 33.66 | 8.95E+02 | 4.48E+01 | 39.0 | Negative |
| PBS 40 | 33.97 | 7.06E+02 | 3.53E+01 | 39.3 | Negative |
| PBS 24 | 34.02 | 6.79E+02 | 3.40E+01 | 39.3 | Negative |
| PBS 56 | 34.14 | 6.19E+02 | 3.10E+01 | 39.4 | Negative |
| PBS 13 | 34.21 | 5.87E+02 | 2.93E+01 | 39.5 | Negative |
| PBS 4 | 36 | 1.49E+02 | 7.43E+00 | 41.3 | Negative |
| PBS 20 | 37.73 | 3.94E+01 | 1.97E+00 | 43.0 | Negative |
| PBS 6 | 37.97 | 3.27E+01 | 1.64E+00 | 43.3 | Negative |

**Figure 1.**
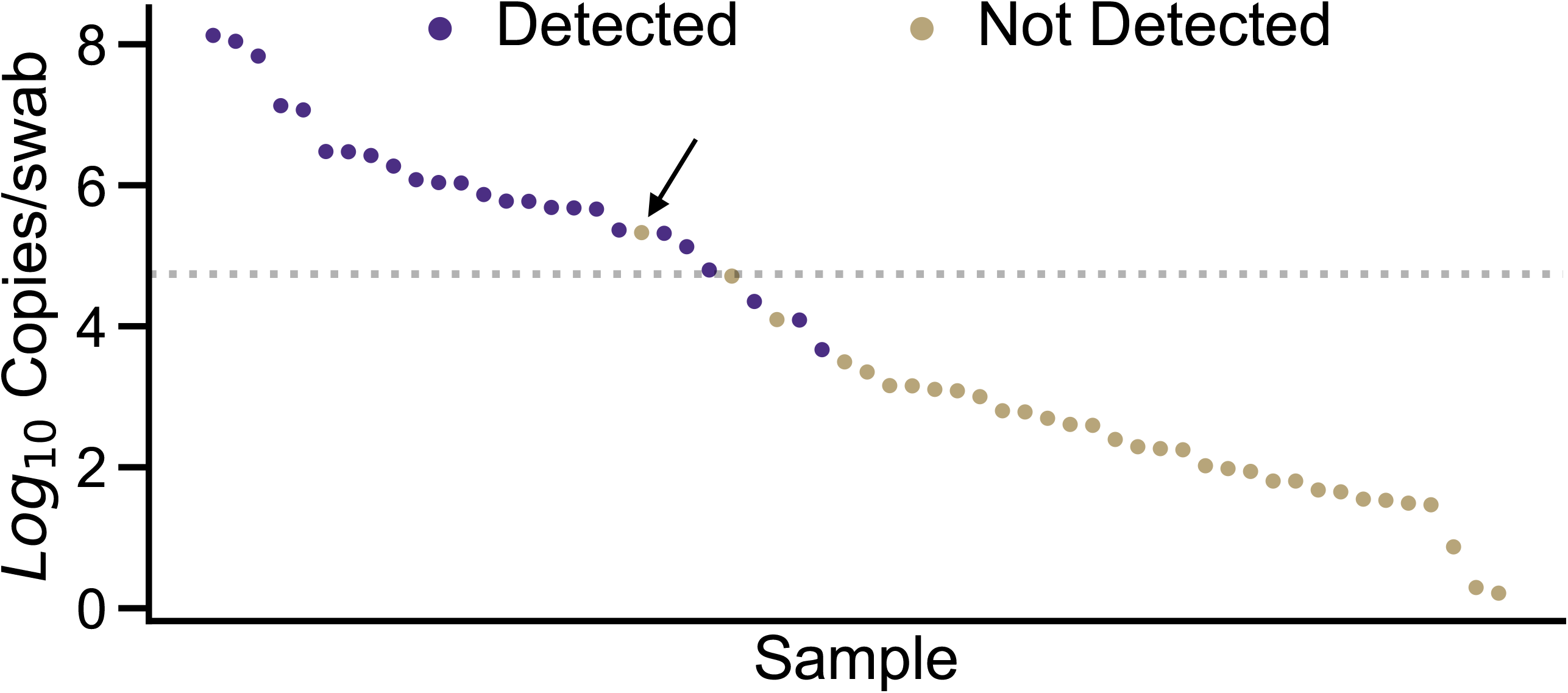
Confirmation of analytical sensitivity of the using clinical specimens. Specimens are depicted in descending order by copies/swab from left to right. Specimens depicted with purple circles were detected by the Quidel Sofia 2 while gold circles were undetected. The analytical sensitivity determined using dilutions of a pooled positive of 54,900 copies/swab is denoted by the dotted line. The arrow denotes the specimen that was negative on repeat testing and subjected to whole genome sequencing.

### Clinical specimens with nucleocapsid T205I/D399N mutation repeatedly test negative on Quidel Sofia 2 antigen test

Although the number of positive detections from blinded SARS-CoV-2 samples within the established LoD was >95%, one sample with a viral load of 213,000 copies/swab – far exceeding the estimated LoD of 48,100-61,100 copies/swab – tested negative on the Quidel Sofia 2. The sample remained negative upon repeat testing on the platform. This sample was re-quantified on a separate molecular platform, the Panther Fusion (Hologic, Marlborough, MA, USA), and was confirmed to contain approximately 2×10^5^ virus copies/swab.

Because of the concern for variants on determining diagnostic assay performance, we performed SARS-CoV-2 whole genome sequencing on this unexpectedly negative specimen. Whole genome sequencing revealed two coding mutations in the N gene, T205I and D399N. Both mutations are located in areas of the 419 aa protein without an available structure, with the T205I in the serine-arginine rich region and the D399N in the far C-terminus (21, 22). Of the 1,144,036 GISAID consensus sequences available as of April 18, 2021, we found 222 sequences with the D399N mutation, for a global prevalence of only 0.019%. This mutation is found mostly in the United States (n=125, 56.3% of total D399N variants), and most commonly in the background of the B.1.429 / 20C clade (n=63) (Figure 2A/B). We further analyzed D399N in the context of other N gene mutations (Figure 3). D399N rarely emerges alone as a single variant in the N gene (n=11, 5.0%), and most frequently appears with only T205I (n=78, 35.1%). T205I is a common N gene mutation in global consensus sequences, at 42.9% prevalence, making the co-occurrence of D399N and T205I a relatively rare event at only 0.16% of total T205I mutations.

**Figure 2.**
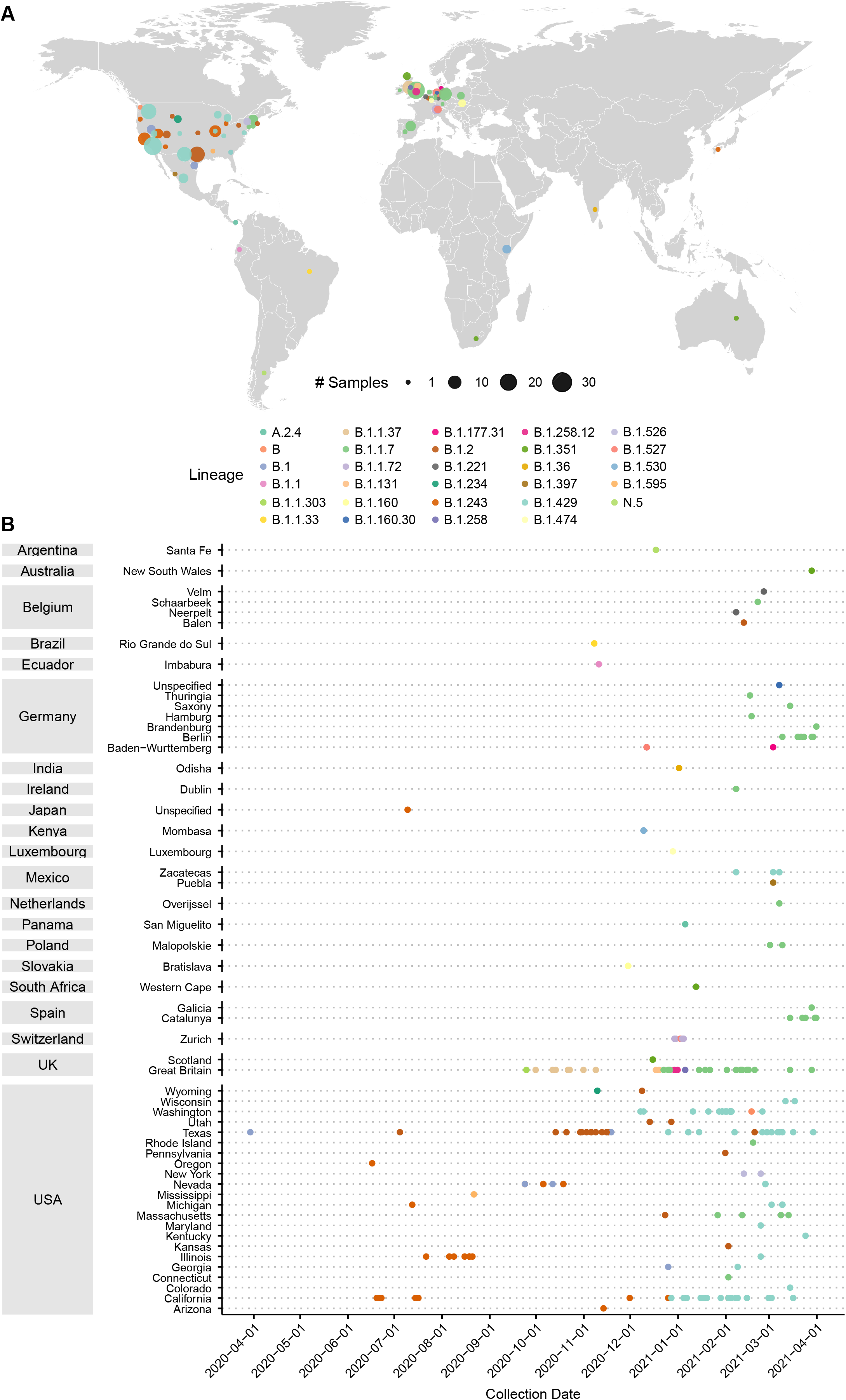
Prevalence of D399N mutations in deposited GISAID consensus sequences. A) Distribution of deposited GISAID genomes with the D399N mutation across the globe. Each dot represents sequences in a GISAID-defined subregion, with area of the dot proportional to the number of sequences. Dots are colored by PANGO lineage. B) Distribution of deposited GISAID genomes with the D399N mutation over time. Countries and subregions are indicated on the left. Each dot represents a unique deposited sequence, colored by PANGO lineage.

**Figure 3.**
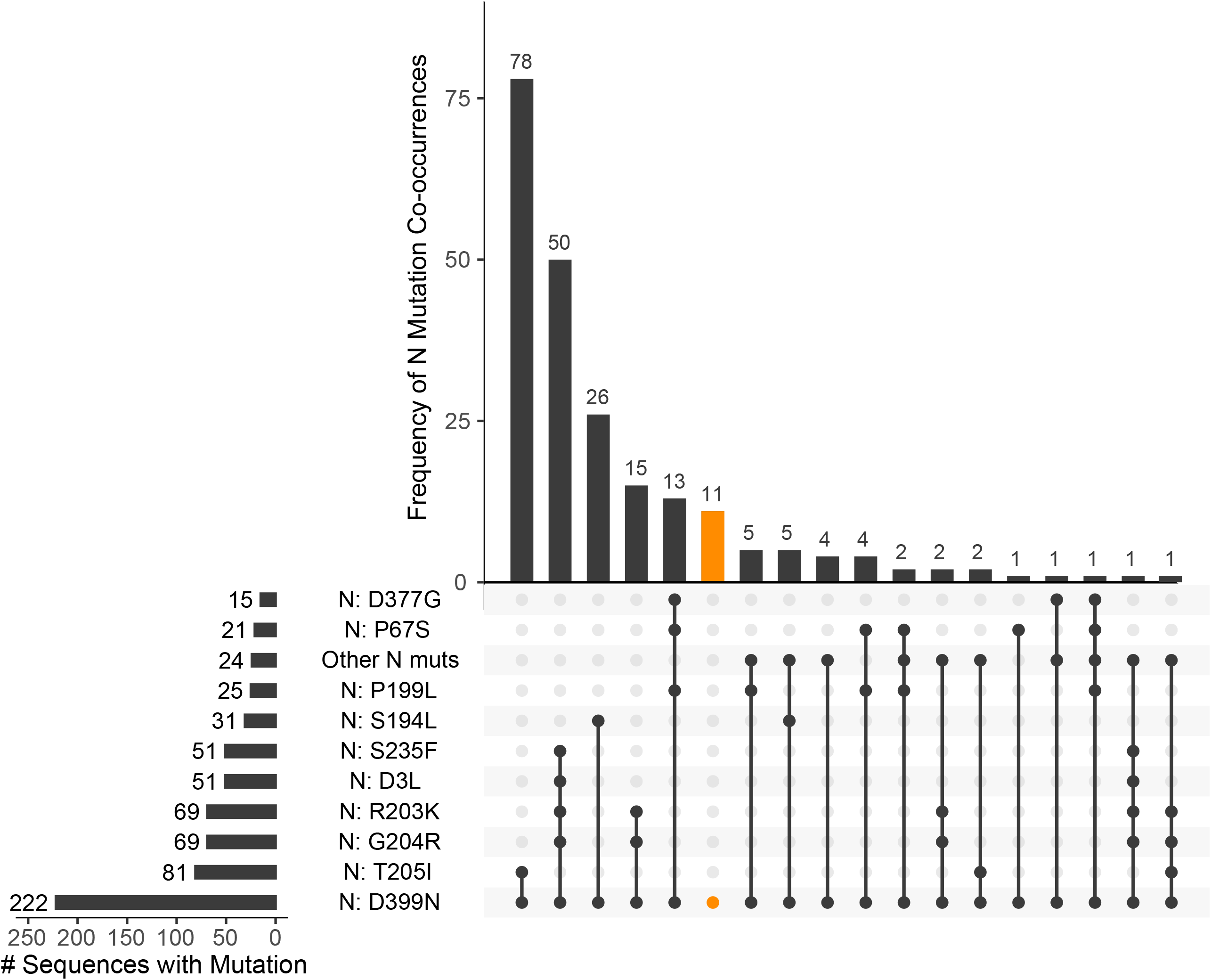
D399N in the context of other N gene mutations in deposited GISAID consensus sequences. Matrix shows co-occurrences of D399N with other N gene mutations, sorted in descending frequency from left to right, with frequencies plotted above as a bar plot. Frequency of each individual mutation is plotted to the left. Highlighted in orange are the genomes with D399N without any co-occurring N gene mutations. N gene mutations with <10 total co-occurrences are collapsed into an “Other N mutations” category.

Upon review of specimens previously sequenced in our lab, we found an additional 6 specimens that contained the same two nucleocapsid mutations and were available for further testing. While 4/6 samples were within the LoD of the Sofia antigen FIA established here, 0/6 samples were detected by the assay. Five of the 6 samples harboring the two N gene mutations were detected on a second antigen test, the Abbott BinaxNOW COVID-19 card. The sixth sample was determined to have a theoretical C_T_ of 37.6, far exceeding the analytical sensitivity of the Abbott BinaxNOW COVID-19, which was previously determined to be an approximate C_T_ of 29-30 (15).

### Ectopically expressed nucleocapsid protein containing the D399N mutation results in a ∼1000-fold reduction in analytical sensitivity specifically on the Quidel Sofia SARS Antigen FIA test

Testing of available clinical samples showed that the Quidel Sofia 2 antigen test has reduced sensitivity for specimens associated with the N T205I/D399N variant, but not for specimens associated with the N T205I variant. Given that the N D399N mutation rarely emerges alone, no clinical specimens were available for antigen testing to directly address whether D399N mutation is solely responsible for the reduced sensitivity of the Quidel Sofia 2 antigen test. To evaluate the role of the D399N mutation alone or in combination with T205I, we transiently expressed N variants in 293T cells and subjected lysates from these 293T cells to antigen testing.

The N wildtype gene from Wuhan-Hu-1 (N WT) and N gene variants N T205I, N D399N, and N T205I/D399N were each cloned into a CMV expression vector with a C-terminal 2XStrep-Tag II. Each of these constructs and the empty vector were transfected into 293T cells and lysates were harvested 48 h post-transfection. Western blot analysis with anti-N of cell lysates and a commercial recombinant N standard was performed to confirm the expression of all recombinant N proteins and to estimate the amount of N protein produced. Based on the estimated nanograms of N loaded into the western blot, the concentration of N in each lysate was estimated to vary from 190 ng/µL to 404 ng/µL (Figure 4).

**Figure 4.**
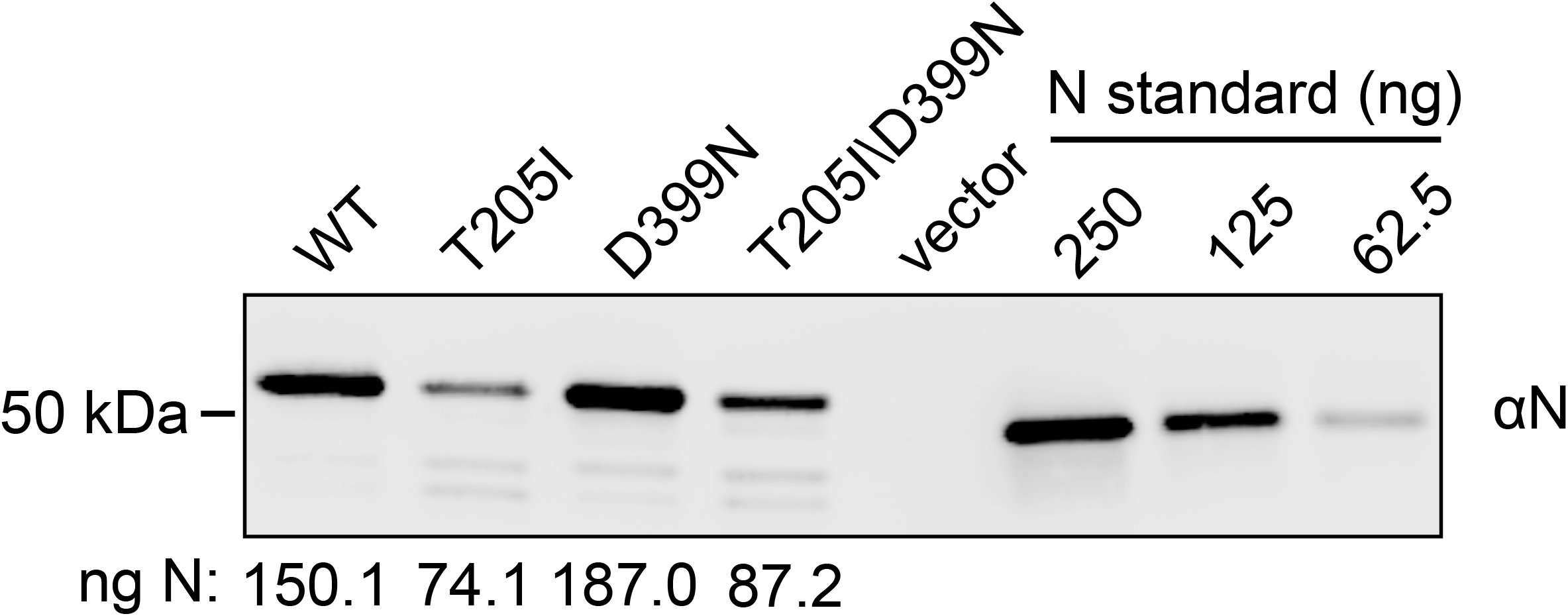
Western blot quantitation of ectopically expressed recombinant SARS-CoV-2 nucleocapsid protein in 293T cells. To evaluate N protein expression, total protein normalized lysates from 293T cells transfected with N Wuhan-Hu-1 (WT), N T205I, N D399N, N T205I/D399M, or empty vector and either 250 ng, 125 ng, or 62.5 ng of recombinant N were subjected to SDS-PAGE followed by □N western blot. The estimated nanograms of N loaded per lane for each variant is shown beneath the western blot determined from a standard curve generated by quantification of the commercial recombinant N standards.

Next, we wanted to compare the analytical sensitivity of the Quidel Sofia 2 and the Abbott BinaxNOW antigen test kits for the N variants expressed in 293T cells. We confirmed that lysates from 293T cells transfected with the empty vector, which did not express N, were not detected (Table 4). To compare the N variants, we first diluted the lysates that contained a higher concentration of N protein by 3-fold and then each lysate was subjected to a 10-fold serial dilution beginning at 1:10 and ending at 1:10^6^ to determine the limit of detection for each antigen test. Based on the western blot quantification, the expected pg of N per swab from the 1:10 dilution is 5.6×10^5^ pg, 9.5×10^5^ pg, 6.7×10^5^ pg, 12.2×10^5^ pg for N WT, N T205I, N D399N, and N T205I/D399N, respectively. In testing of clinical specimens, as described above, the Abbott BinaxNOW test detected specimens associated with both N T205I and N T205I/D399N variants with similar sensitivity. Likewise, the Abbott BinaxNOW kit detected each of these N variants in 293T lysates with equivalent sensitivity down to the 1:10^5^ dilution (3/3 or 2/3 positive tests), corresponding to approximately 95 pg of N205I and 122 pg of N T205I/D399N (Table 4). Furthermore, the Abbott BinaxNOW test detected lysates that contain N with just the single mutation D399N or N WT with sensitivity equivalent to the other N variants. Thus, the Abbott BinaxNOW test appears unaffected by the N variants T205I, D399N, or T205/D399N.

**Table 3.** Antigen test results for available clinical specimens containing D399N and T205I nucleocapsid mutants

| Specimen | Cobas 6800 C<sub>T</sub><br>(E-gene) | Theoretical<br>Roche cobas<br>C<sub>T</sub> | Quidel<br>Sofia 2 | Abbott<br>BinaxNOW | Nucleocapsid<br>Mutation | Lineage | GISAID EPI |
| --- | --- | --- | --- | --- | --- | --- | --- |
| DM1 | 32.58 | 37.9 | Negative | Negative | T205I, D399N | B.1.429 | EPI_ISL_1209060 |
| DM2 | 21.85 | 27.2 | Negative | Positive | T205I, D399N | B.1.429 | EPI_ISL_1110097 |
| DM3 | 25.48 | 30.8 | Negative | Positive | T205I, D399N | B.1.429 | EPI_ISL_1789833 |
| DM4 | 20.44 | 25.8 | Negative | Positive | T205I, D399N | B.1.429 | EPI_ISL_1069365 |
| DM5 | 21.72 | 27.1 | Negative | Positive | T205I, D399N | B.1.429 | EPI_ISL_1366181 |
| DM6 | 23.25 | 28.6 | Negative | Positive | T205I, D399N | B.1.429 | EPI_ISL_1366195 |
| DM7* | 22.63 | 28.0 | Negative | n.d. | T205I, D399N | B.1.429 | EPI_ISL_1708574 |
| M8 | 15.29 | 20.7 | Positive | Positive | T205I | B.1.429 | EPI_ISL_1405087 |
| M9** | 24.27 | 29.6 | Positive | Negative | T205I | B.1.429 | EPI_ISL_1405089 |
| M10** | 22.3 | 27.6 | Negative | Positive | T205I | B.1.429 | EPI_ISL_1405090 |
| M11 | 21.21 | 26.6 | Positive | Positive | T205I | B.1.429 | EPI_ISL_1405112 |
| M12 | 24.41 | 29.7 | Positive | Positive | T205I | B.1.429 | EPI_ISL_1405113 |
| M13 | 22.47 | 27.8 | Positive | Positive | T205I | B.1.429 | EPI_ISL_1405115 |
| M14 | 24.06 | 29.4 | Positive | Positive | T205I | B.1.429 | EPI_ISL_1405119 |
| M15 | 23.4 | 28.7 | Positive | Positive | T205I | B.1.429 | EPI_ISL_1405138 |
| M16 | 20.06 | 25.4 | Positive | Positive | T205I | B.1.429 | EPI_ISL_1405141 |
| M17 | 22.78 | 28.1 | Positive | Positive | T205I | B.1.429 | EPI_ISL_1405148 |
| M18 | 17.47 | 22.8 | Positive | Positive | T205I | B.1.429 | EPI_ISL_1405151 |

\*Original discordant sample
\*\*M9 and M10 were diluted to create enough volume to run on cobas 6800 for a fresh C<sub>T</sub> (Dilution was prepared prior to both antigen and molecular testing)
n.d., not done

**Table 4.** Antigen testing results for detection of transiently expressed nucleocapsid proteins. The approximate initial limit of detection is highlighted in bold, where both highlighted 2/3 decisions involved a split decision on the 3^rd^ specimen in question. The estimated amount of protein placed on a swab was calculated based on the western blot in Figure 4.

| Sample | Dilution | Sofia 2 | BinaxNOW | QuickVue | Estimated protein (pg) |
| --- | --- | --- | --- | --- | --- |
| Negative Control | 1:100 | 0/1 | 0/1 | 0/3 |  |
| Wild-type N | 1:10 | 3/3 |  | 3/3 | 560400 |
|  | 1:100 | 3/3 |  | 3/3 | 56040 |
|  | 1:1000 | 3/3 | 3/3 | 3/3 | 5604 |
|  | 1:10000 | **3/3** | 3/3 | **3/3** | 560 |
|  | 1:100000 | 0/3 | **3/3** | **2/3** | 56 |
|  | 1:1000000 |  | 0/3 | 0/3 | 6 |
| N T205I | 1:10 | 3/3 | 1/1 | 1/1 | 948500 |
|  | 1:100 | 3/3 | 1/1 | 3/3 | 94850 |
|  | 1:1000 | 3/3 | 3/3 | 3/3 | 9485 |
|  | 1:10000 | **3/3** | **3/3** | 3/3 | 948 |
|  | 1:100000 | 0/3 | **2/3** | **3/3** | 95 |
|  | 1:1000000 |  | 0/3 | 0/3 | 9 |
| N D399N | 1:10 | **6/6** |  | 3/3 | 673200 |
|  | 1:100 | 1/6 |  | 3/3 | 67320 |
|  | 1:1000 | 0/3 | 3/3 | 3/3 | 6732 |
|  | 1:10000 | 0/3 | 3/3 | **3/3** | 673 |
|  | 1:100000 | 0/3 | **3/3** | **2/3** | 67 |
|  | 1:1000000 |  | 0/3 | 0/3 | 7 |
| N T205I/D399N | 1:10 | **4/4** | 1/1 | 1/1 | 1220800 |
|  | 1:100 | 2/6 | 1/1 | 3/3 | 122100 |
|  | 1:1000 | 0/3 | 3/3 | 3/3 | 12210 |
|  | 1:10000 | 0/3 | 3/3 | 3/3 | 1221 |
|  | 1:100000 | 0/3 | **3/3** | **3/3** | 122 |
|  | 1:1000000 |  | 0/3 | 0/3 | 12 |

In contrast to the Abbott BinaxNOW kit, the Quidel Sofia 2 antigen test exhibited significantly lower sensitivity for clinical specimens associated with N T205I/D399N compared to clinical specimens associated with N T205I. Testing of 293T lysates containing N T205I or N T205I/D399N on the Quidel Sofia 2 antigen test confirmed this finding with the antigen test exhibiting an approximately 1000-fold reduced sensitivity between lysates containing variants N T205I versus N T205I/D399N, based on comparing the final dilution where all tests were positive. This corresponded to a difference of approximately 948 pg N T205I and 1.22×10^6^ pg of N T205I/D399N (Table 4). In contrast, Quidel Sofia 2 antigen test has approximately equivalent sensitivity between lysates containing N WT and N T205I since both were 100% detected (3/3) at 1:10^4^ dilution or approximately 560 pg of N WT protein and 948 pg of N T205I protein.

To determine if D399N alone is sufficient for the reduced sensitivity of the Quidel Sofia 2 antigen test, we next tested lysates that contain N D399N mutation alone. In contrast to lysates containing N T205I alone or N WT, the sensitivity of the Quidel Sofia 2 antigen test for lysates containing N D399N mutation alone was reduced by 1000-fold (comparing the last dilution where all tests were positive), similar to lysates containing the N T205I/D399N double mutant. (Table 4). These findings demonstrate that the N D399N mutation alone was sufficient to reduce the sensitivity of the Quidel Sofia 2 antigen test significantly by 1000-fold.

We next tested the 293T lysates containing N variants on another point-of-care, rapid, FDA authorized COVID-19 antigen test manufactured by Quidel Corporation, Quidel QuickVue, to determine if the Quidel QuickVue kit sensitivity is affected by the N D399N mutation. The QuickVue test is performed as a dipstick test and does not require a reader like the Sofia 2 test. In contrast to our findings on the Quidel Sofia 2 antigen tests, no significant difference was observed in sensitivity between any of the N variants, indicating that the Quidel QuickVue test is not affected by the D399N mutation (Table 4).

## Discussion

Here, we simultaneously determined the analytical sensitivity of the Quidel Sofia SARS Antigen FIA run on the Sofia 2 analyzer and uncovered a nucleocapsid variant that dramatically affects the analytical sensitivity of this antigen test. We found the analytical sensitivity of the Quidel Sofia 2 corresponded to 48,100 – 61,100 copies/swab, which closely matches the analytical sensitivity (40,400 – 80,600 copies/swab) we found for the Abbott BinaxNOW (15). While not absolutely comparable, the relative analytical sensitivities agree with those listed in the package inserts for the Quidel Sofia 2 and Abbott BinaxNOW assays of 113 TCID_50_ units/mL and 140.6 TCID_50_ units/mL, respectively. Our limited testing against recombinant proteins hinted at a slightly better analytical sensitivity for the Abbott BinaxNOW and Quickvue assays compared to the Quidel Sofia SARS Antigen FIA. However, this testing was run against comparatively few samples and mainly used to demonstrate the semi-quantitative effects of nucleocapsid mutations on test performance.

We further characterized a single amino acid mutation (D399N) that affected the analytical sensitivity of the Quidel Sofia SARS Antigen FIA by approximately 1000-fold, when measured on the Sofia 2. The D399N mutation that affected the Quidel Sofia 2 assay performance seems to be of limited public health importance at this time given the few number of cases it has been detected in. It is also a mystery as to how this amino acid change could affect nucleocapsid function. Of note, no complete SARS-CoV-2 nucleocapsid structures are available, with only the N-terminal RNA-binding region and the C-terminal domain (21, 22). Neither the T205I or D399N mutation recovered in our interrogated specimens occurred in these regions, nor could high-confidence models be created for these regions in HHPred (23). Given that one of these mutations affected diagnostic sensitivity, our data illustrate the comparable lack of understanding of how mutations can affect coronavirus nucleocapsid protein as compared to the spike protein.

Compared to molecular testing, antigen test performance is often less well-characterized and the effect of viral variation on assay performance only compounds this issue. Antigen tests are often performed in a distributed manner, away from the availability of comparator platforms to monitor test performance. Antigen tests often make use of the entire specimen, obviating the ability to run same sample comparisons on other platforms. And, while it is relatively easy to understand how viral mutation could affect PCR primer binding – even if they are not routinely publicly available for most FDA authorized platforms – antigen testing is often literally a black box, with no public epitope mapping information or even knowledge of whether such data exists. Increasingly, initiatives from the NIH, CDC, and FDA are trying to systematically examine the effect of variants on diagnostic performance, but it is worth noting that multiple molecular tests have already been found to be affected by variants while the first antigen test affected has not been described until more than a year after authorization (24, 25).

The main limitations of the study include the small number of specimens tested, use of specimens that do not follow the manufacturer’s recommended testing protocol, use of unpurified recombinant protein lysates to confirm mutation effects, and limited testing to initial determination of analytical sensitivity using recombinant protein. Nonetheless, we found consistent results between use of clinical specimens and recombinant proteins that bulwarks our results. The Quidel Sofia SARS Antigen FIA is a sandwich ELISA which involves both a capture and detection antibody. At this time, it is not clear whether the D399N mutation alters binding by the capture or the detection antibody. Without the antibodies used in these assays, it is difficult to determine equilibrium dissociation constants, perform epitope mapping, or measure other biochemical properties associated with the test reagents.

Our data further highlight the increasing returns of widespread genome sequencing in the clinical microbiology community (26, 27). With routine SARS-CoV-2 whole genome sequencing available in our lab, we were rapidly able to determine the coding mutations present in a diagnostic edge case. This mutation could further be probed by examining more specimens for which routine genome sequencing had been performed. All told, the work in the clinical virology described here was completed in under four weeks and the antigen test manufacturer was made aware within four days of our testing of the genotyped clinical specimens. Although this specific story uncovered a variant that affected SARS-CoV-2 antigen testing, the ability of decentralized, widespread, routine whole genome sequencing to uncover the genetic basis of assay performance could equally be applied to almost any diagnostic test in the clinical microbiology lab.

## Data Availability

Testing data available in tables and from GISAID.

## Acknowledgements

The authors thank the staff of UW Virology as well as financial support from the Chisholm Foundation, and Quidel and Abbott for providing in-kind kits for testing. The funders had no role in study design, data collection and interpretation, or the decision to submit the work for publication. ALG reports contract testing from Abbott and research support from Merck and Gilead. The authors also gratefully acknowledge authors from originating laboratories responsible for obtaining the specimens and the submitting laboratories where genetic sequence data were generated and shared via the GISAID Initiative, on which some of this research is based.

